# Common and rare variants in *SLCO1B1* are associated with statin intolerance

**DOI:** 10.1101/2022.08.09.22278584

**Authors:** Margherita Bigossi, Cyrielle Maroteau, Adem Y Dawed, Alasdair Taylor, Sundararajan Srinivasan, Alaa’ Lufti Melhem, Ewan R Pearson, Roberto Pola, Colin N A Palmer, Moneeza K Siddiqui

## Abstract

**Background and Aims:** The efficacy of statin therapy is hindered by adverse drug reactions, most frequently musculoskeletal symptoms. Variants in *SLCO1B1*, which encodes the hepatic transporter OATB1B1, influence statin pharmacokinetics, resulting in altered plasma concentration of the drug and its metabolites. While pharmacogenetic testing of the loss-of-function Val174Ala (rs4149056T>C) is recommended to reduce risk of statin intolerance, current guidelines acknowledge the potential role of gain-of-function variants. This study tests the hypothesis that accounting for gain-of-function variants in *SLCO1B1*, in addition to Val174Ala, will provide more reliable estimates of statin intolerance.

**Methods and Results:** High-risk haplotypes were derived from Val174Ala and three common gain-of-function *SLCO1B1* variants and compared to low-risk haplotypes. In statin users from Tayside Scotland, UK, those with high-risk haplotypes had increased odds across three phenotypes of statin intolerance (general statin intolerance: OR_GSI_ 2.42[95%CI:1.29, 4.31], p=0.003; statin-related myopathy OR_SRM_ 2.51[95%CI:1.28, 4.53],p=0.004; statin-related suspected rhabdomyolysis: OR_SRSR_ 2.85[95%CI:1.03, 6.65],p=0.02). In contrast, using the Val174Ala genotype alone produced weaker results. A meta-analysis with results from adjudicated cases of statin-induced myopathy in the PREDICTION-ADR Consortium confirmed these findings (OR_Val174Ala_ 1.99 [95%CI:1.01, 3.95],p=0.048; OR_risk-haplotypes_ 1.76 [95%CI:1.16, 2.69],p=0.008). For those requiring high-dose statin therapy, high-risk haplotypes were more consistently associated with the time to onset of statin intolerance amongst the three phenotypes compared to Val174Ala (general statin intolerance: HR_Val174Ala_ 2.49 [95%CI:1.09, 5.68],p=0.03; HR_risk-haplotypes_ 2.44 [95%CI:1.46, 4.08],p<0.001). Exome-sequenced rare variants were found to be associated with the risk of intolerance (p=0.02).

**Conclusions:** We demonstrate that accounting for gain-of-function variants in *SLCO1B1*, in addition to Val174Ala, provides more reliable estimates of statin intolerance.

## Introduction

Statins are the cornerstone of lipid-lowering therapy, showing great efficacy in reducing cholesterol levels and the incidence of cardiovascular events. Evidence from multiple randomized clinical trials (RCT) and meta-analyses show that statin therapy reduces major cardiovascular events (MACE) by ∼22% for each mmol/L reduction in LDL-cholesterol (LDL-C), and total mortality by ∼10% over 5 years. Adverse drug reactions (ADRs) to statins in clinical trials were rare, presenting in only ∼5% of cases^1^. Despite this favourable safety profile, in routine care settings nearly one in five patients on statin therapy discontinues the drug due to reported side effects^2^. Musculoskeletal symptoms are the most common ADRs. These can range from muscle pain or weakness associated with increased creatinine kinase levels (i.e., myalgia and myopathy), to rapid and life-threatening muscle breakdown (i.e., rhabdomyolysis). These symptoms are the primary cause of poor compliance to statin therapy, accounting for an estimated 25% relative risk increase in the rate of first-onset cardiovascular events^3^.

The causal mechanisms of these side effects are not fully understood; however, there is evidence of a correlation with systemic exposure to statins and their metabolites. Key enzymes in the pharmacokinetics of statins are those responsible for their uptake, metabolism, and elimination. The most extensively studied protein in relation to statin intolerance is the hepatic uptake transporter OATP1B1. Missense variants in the encoding gene, *SLCO1B1*, are associated with altered functional activity of the transporter. This is consequently linked with the systemic concentration of statins and their metabolites, and is therefore associated with the risk of statin intolerance. The strongest evidence exists for a common loss-of-function (LoF) variant, rs4149056:Val174Ala. Homozygous carriers of the minor C allele at rs4149056 (*5/*5) show increased plasma concentrations of statins by as much as 221% in *in vitro* studies, and an increase in the odds of myopathy by 3-to-17-fold in post-hoc analyses of RCTs^4-11^.

The effect of other *SLCO1B1* variants on statin-associated myopathy is less clear. In a recent GWAS of simvastatin metabolites by Mykkänen et al., two gain-of-function (GoF) variants (rs11045819:Pro155Thr and rs34671512:Leu643Phe) were independently associated with a 32% and 36% per allele decrease in systemic exposure to simvastatin acid, respectively^12^. These pharmacokinetic findings support the hypothesis that while Val174Ala is exerting a loss of function effect, there are independent GoF variants in *SLCO1B1*. Indeed, a study by Ramsey et al., focused on the effect of all non-synonymous (NS) variants in *SLCO1B1* in methotrexate clearance, confirmed the independent role of these variants ^13^. Among the 15 variants studied only 4 were commonly occurring (MAF >5% in the general population). The only additional variant identified by Ramsay et al. was the GoF rs2306283:Asn130Asp, which in conjunction with Val174Ala has been shown to have a role in statin response and ADR^13,14^. Common variants in *SLCO1B1* accounted for 8.72% of the population variability in methotrexate clearance, while rare variants accounted for 2% of the variability ^13^.

Current CPIC recommendations for the management of statin-induced myopathy are centered around the *SLCO1B1* Val174Ala variant while acknowledging the potential role of GoF ^15,16^. The strongest evidence for recommendations is for simvastatin and atorvastatin users. The new functional phenotypes of *SLCO1B1* identify differentially decreased OATP1B1 function between heterozygous and homozygous carriers of Val174Ala. Taken together with functional studies in methotrexate, this suggests a potential role for GoF variants in explaining the lower risk for heterozygous carriers of Val174Ala.

In this study, we use large cohorts to test the hypothesis that accounting for GoF variants in *SLCO1B1*, in addition to Val174Ala, results in more reliable associations with statin intolerance in a real-world setting. Additionally, we use whole-exome sequencing to estimate the role of rare *SLCO1B1* variants in the development of clinically adjudicated statin-induced myopathy.

## Methods

This paper has been reported according to the STREGA (STrengthening the REporting of Genetic Association Studies) guidelines.

### Discovery Cohort: Tayside Bioresource

This case-control study population was derived from two bioresources: GoDARTS (Genetics of Diabetes Audit and Research in Tayside Scotland) and SHARE (Scottish Health Research Register and Biobank), which are part of the Tayside Bioresource, University of Dundee ^17,18^. Details on recruitment, data collections, linkage, and biobanking have been described in detail previously^17,18^. These bioresources contain electronic health records, genetic data, laboratory information and community prescribing records with longitudinal follow-up from a total of 15,378 statin users of White European descent. These bioresources have been previously used to make discoveries in the pharmacogenetics of statins, blood pressure medications and anti-diabetic drugs^14,19-22^.

A statin user was defined as an individual with two or more prescriptions of a statin. Historical prescriptions of statins include simvastatin, atorvastatin, rosuvastatin, cerivastatin, pravastatin, fluvastatin. Data were available from January 1999 to December 2020.

#### Phenotypes of statin intolerance

Prescribing patterns and laboratory testing data were used to establish phenotypes of statin intolerance. Statin types, average daily dose, statin switching, discontinuation and percent daily coverage were computed from prescribing records. The use of prescribing patterns to identify statin intolerance has been described previously^20^. CK test measurements from outpatient or inpatient settings were collected. CK tests from high dependency units (Emergency Rooms, Cardiac Care, Stroke, Trauma Units or surgical wards) were excluded. The upper limit of normal (ULN) for usable test results was set at 120 IU/L for women and 180 IU/L for men, based on criteria used and described previously^23^. The highest CK test results while on statin per individual were used to define intolerance. Three phenotypes of increasing severity of intolerance were defined in order to assess for a biological gradient of genetic effect.

##### General Statin Intolerance (GSI)

Cases of general statin intolerance (GSI) were defined as users with at least one CK measurement above the ULN while on statin therapy, associated to either three or more switches in statin drug or premature discontinuation of treatment (defined as interruption of treatment 9 months from the date of death or from the study ending date, whichever came first). The definition of GSI has been published as part of the discovery of the role of the *LILRB5* variant in statin intolerance^20^.

##### Statin-Related Myopathy (SRM) and Statin-Related Suspected Rhabdomyolysis (SRSR)

Cases of statin-related myopathy (SRM) were defined as users with at least one CK measurement 4 times above the ULN while on statin therapy. Statin users with at least one CK measurement 10 times above the ULN were considered to be possible cases of statin-related rhabdomyolysis (SRSR).

##### Statin Tolerance (ST)

The definition of statin tolerance has been defined and used previously^20^. Briefly, controls were defined as statin users who had been on the drug for at least 5 years with adherence of at least 90% and a minimum average daily dose of 40 mg (calculated in simvastatin equivalents)^24^, and had to have no recorded high CK measurement, no discontinuation, and no more than one switch in statin therapy.

##### Validation of phenotypes

These definitions of statin intolerance were validated against the outcome of major adverse cardiovascular events. Hazards of MACE were significantly higher for those with intolerance compared to those who were tolerant of their statins. GSI was associated with 1.39 times the hazards of MACE, while SRM and SIR were associated with 2.18 and 3.06 times the hazards of MACE compared to statin tolerant controls. For details on MACE-free survival analysis see Supplementary methods (Supplementary 3: Validation of phenotypes of intolerance).

#### Genetic data for common variants

Genotype data for the 4 common *SLCO1B1* polymorphisms (rs4149056:Val174Ala, rs2306283:Asn130Asp, rs11045819:Pro155Thr, rs34671512:Leu643Phe) were available from six platforms. Details on genotyping methods are available in Supplementary methods (Supplementary methods 1: Genetic data platforms). The genetic quality control methods used for this cohort have been described previously^17^. Genotype quality threshold of 90% was applied for imputed SNPs. Minor allele frequencies for each variant were similar to those of a reference white European population, and their genotype frequencies did not deviate from Hardy-Weinberg equilibrium (details in Supplementary methods, Table 1).

**Table 1.** Definition of the high-risk haplotypes from the four common *SLCO1B1* variants

|  | **Val174Ala**<br>**(rs4149056)** | **Leu643Phe**<br>**(rs34671512)** | **Asn130Asp**<br>**(rs2306283)** | **Pro155Thr**<br>**(rs11045819)** |
| --- | --- | --- | --- | --- |
| **\*5/\*5** | Ala174Ala | X | X | X |
| **\*1a/\*5** | Val174Ala | Leu643Leu | Asn130Asn | Pro155Pro |

High-risk haplotypes were classified as being either homozygous for the deleterious Val174Ala variant (SLCO1B1 \*5/\*5) or heterozygous for the same variant and not carrying protective variants in the other loci (SLCO1B1 \*1a/\*5). All remaining haplotypes were categorised as low-risk. X indicates either allele at the locus.

#### Defining high-risk haplotypes using common variants

Current pharmacogenetics guidelines classify individuals who are homozygous for Val174Ala (*SLCO1B1*\*5/*5) as being at highest risk of myopathy, while heterozygous carriers for Val174Ala (also referred to as *SLCO1B1*\*5) are at intermediate risk, regardless of the presence of GoF variants. Based on the studies by Ramsay et al. and Mykkänen et al., we defined two high-risk haplotypes using the four common *SLCO1B1* variants (Val174Ala, Leu643Phe, Asn130Asp, Pro155Thr)^12,13^. Similar to CPIC guidelines, those with Ala174Ala (*SLCO1B1*\*5/*5), irrespective of the presence of GoF variants, were considered to have a high-risk haplotype^16^. However, among those heterozygous for Val174Ala (*SLCO1B1*\*5), only those who were not carriers of any GoF variants (*SLCO1B1*\*1a/*5) were considered to have a high-risk haplotype (see Table 1). All remaining haplotypes were categorised as low-risk.

#### Data Source: PREDICTION-ADR and exome sequencing for rare variants

Whole exome-sequencing was undertaken on 229 individuals clinically adjudicated as having statin-induced myopathy and 488 statin tolerant controls as part of the PREDICTION-ADR consortium. Exome sequenced data from this study as well as case adjudication methods have previously been described in the context of discovery and validation of pharmacogenetic association studies^20,21,25^. Details on library preparation, sequencing, read mapping, variant calling, and data quality control have been described previously^20,21^ (details also in Supplementary 2: Exome sequencing for rare variants).

#### Data Source: UK Biobank primary data

The UK Biobank (UKBB) is a large bioresource containing linked electronic medical records and genetic information from 500,000 individuals recruited from 2006 to 2010^26^. In addition to hospital records and genetic data, longitudinal primary care data for around 45% of the UKBB cohort is available.

Phenotype replication was attempted in 61,512 statin users identified from prescription records. In this cohort, longitudinal blood testing results are available from primary care data, available for ∼45% of the UKBB cohort. This resulted in missing data for ∼80% of statin users. Thus, we decided against including UKBB data in further analyses. Further details on the UKBB cohort and CK measurements are available in Supplementary methods (Supplementary 4: UK Biobank cohort).

### Statistical methods

#### Common variants

##### Analyses in Tayside Bioresource

Binary logistic regression was used to test the association between the risk haplotypes (RH) and the phenotypes of statin intolerance. Covariates associated with statin intolerance were used in the models, including sex, age, type of statin drug, average daily dosage and concomitant drug therapy. The relative likelihood AIC statistics were used to compare the established Val174Ala model with the novel risk haplotypes model^27^. Cox proportional-hazards models were used to calculate hazard ratios and 95% confidence intervals for the association of the haplotypes and intolerance-free survival stratified by dose.

##### Meta-analysis with PREDICTION-ADR

Association between *SLCO1B1* (both Val174Ala and risk haplotypes) and clinically adjudicated cases of statin-induced myopathy was tested using a binary logistic regression model adjusted for age and sex. Findings from these analyses were used to perform fixed-effects meta-analyses with results from the Tayside Bioresources. Since the three phenotype groups from Tayside Bioresources contain overlapping individuals, only one of them could be selected for the meta-analysis. The GSI phenotype was selected as it had the largest number of cases. The two meta-analyses were performed using the metafor package in R and results presented in a combined Forest plot.

#### Rare variants

Using exome sequencing data, we performed first a single variant analysis using PLINK/Seq (http://atgu.mgh.harvard.edu/plinkseq/) and then gene-burden testing to identify rare variants associated with clinically adjudicated statin-induced myopathy. Gene-based associations for rare variants (MAF ≤ 1%) were performed using a sequence kernel association test – optimal (SKAT-O)^28^. Methods on sequencing and rare variants analysis have been described in detail previously^21^.

## Results

### Baseline characteristics of the study population

From 15,378 statin users, a total of approximately 174,000 person-years of statin use were available. Approximately 4% of statin users in the study population met the criteria for at least one phenotype of intolerance. Most were cases of GSI (n = 364, 2.4%), followed by cases of SRM (n = 282, 1.8%). As expected, cases of SRSR were the least common (n = 107, 0.7%). 2,309 (15%) statin users met all criteria defining statin tolerant individuals and were used as controls. Baseline characteristics of cases and controls are presented in Table 2.

**Table 2.**
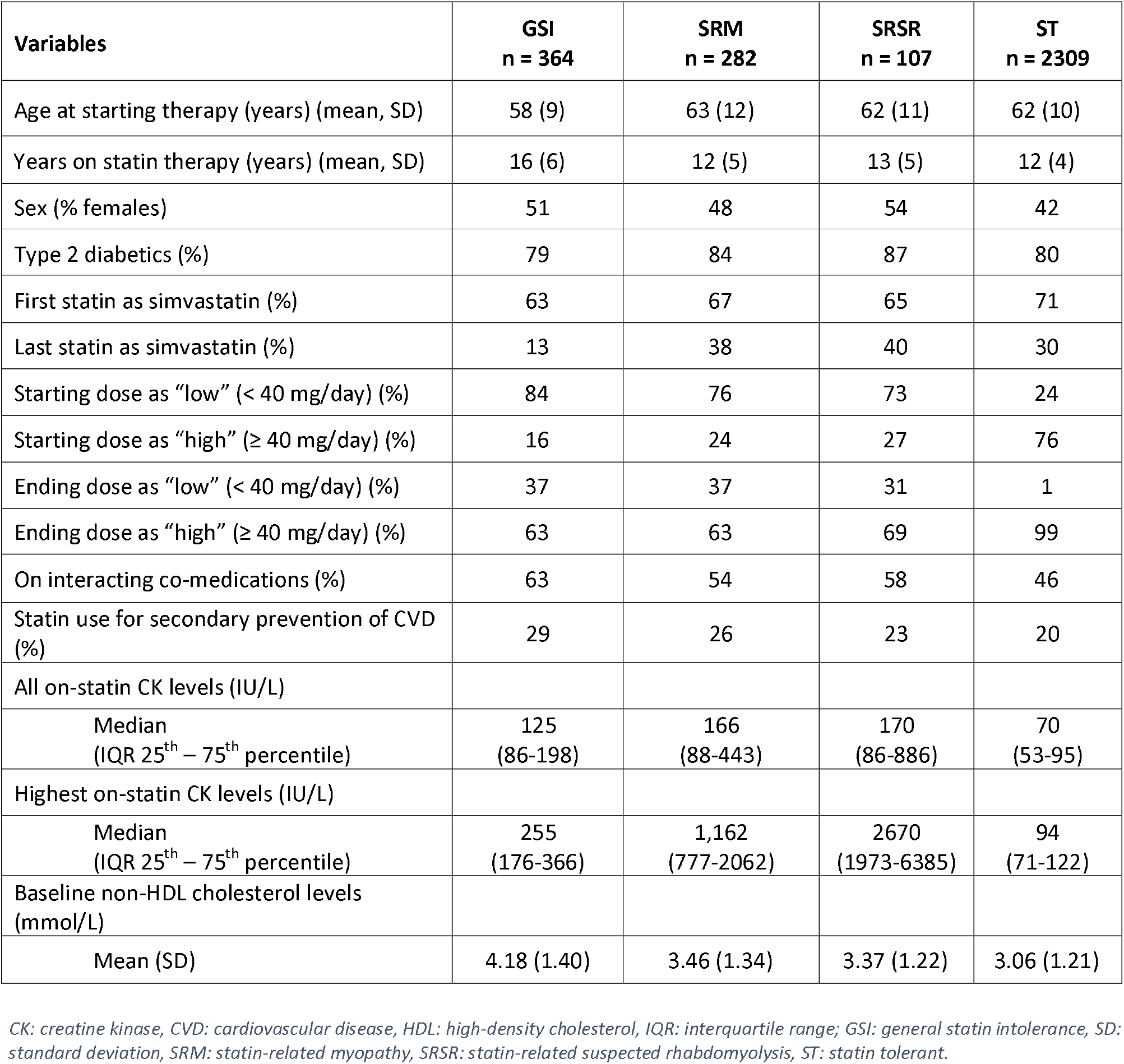
Characteristics of the study population.

### Common variants

#### Comparative association of risk haplotypes and Val174Ala with phenotypes of statin intolerance

Individuals with high-risk haplotypes had increased odds of GSI, SRM and SRSR when compared to those with low-risk haplotypes (GSI: OR 2.42 [95% CI: 1.29, 4.31], p = 0.003; SRM OR 2.51 [95% CI: 1.28, 4.53], p = 0.004; SRSR: OR 2.85 [95% CI: 1.03, 6.65], p = 0.02). In contrast, using the Val174Ala genotype alone produced weaker results. When compared to those with the Val174X genotypes individuals homozygous for Ala174Ala had significantly increased odds of SRM, but not of GSI or SRSR (GSI: OR 2.34 [0.84, 5.62] p = 0.07), SRM: OR 2.71 [95% CI: 1.01, 6.15], p = 0.02; SRSR: OR 2.22 [95% CI: 0.35, 7.8], p = 0.2) (Fig. 2, and corresponding table in Supplementary: Table 3). The AIC criteria for non-nested model selection strongly favoured the haplotypes as the best-fit between the two models (see Supplementary: Table 4).

**Figure 1.**
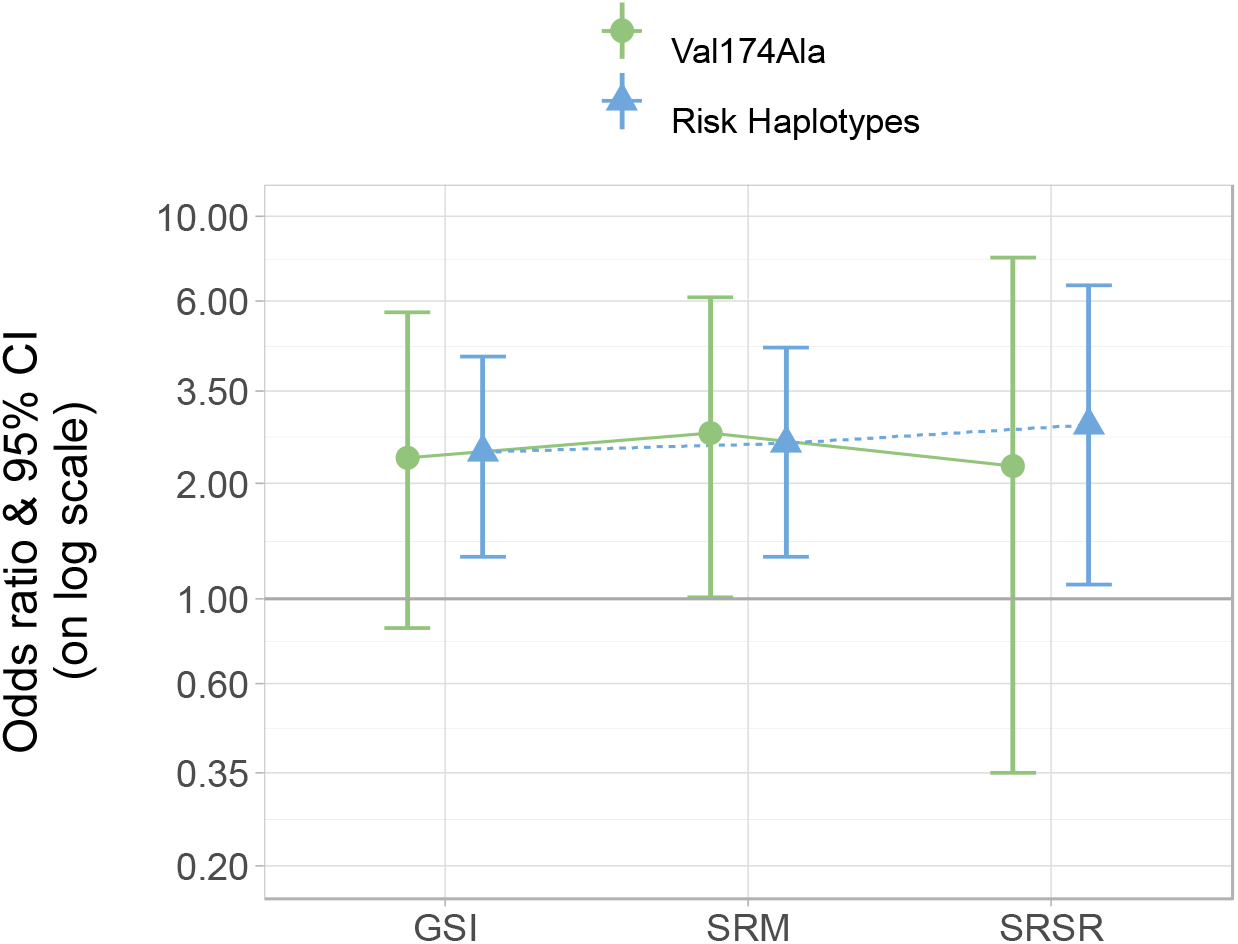
Association of Val174Ala and risk haplotypes with phenotypes of statin intolerance in all statin users. GSI: general statin intolerance, CI: confidence interval, SRM: statin-related myopathy, SRSR: Statin-related suspected rhabdomyolysis. Models adjusted for age, sex, previous cardiovascular events, interacting medications, statin type and statin dose.

**Figure 2.**
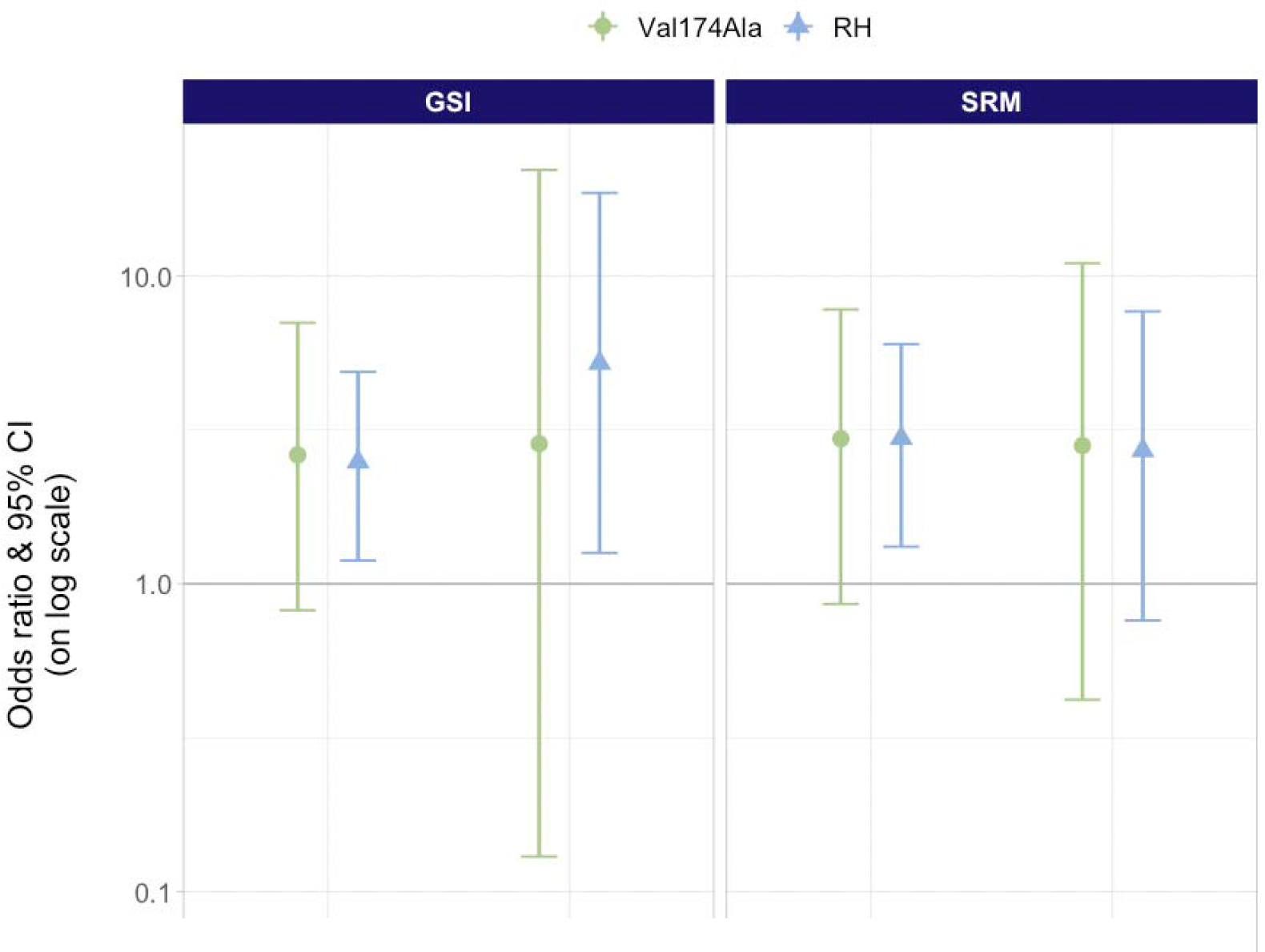
Subgroup analysis in simvastatin and atorvastatin users for the association of Val174Ala and risk haplotypes with phenotypes of statin intolerance. GSI: general statin intolerance, CI: confidence interval, SRM: statin-related myopathy.

#### Risk haplotypes, and not Val174Ala, are associated with intolerance in simvastatin users

Since evidence on the effect of Val174Ala on the risk of intolerance is weaker for drugs other than simvastatin, we performed subgroup analyses to assess the effect of these haplotypes on simvastatin and atorvastatin. Stratifying by first prescribed statin, individuals on simvastatin with high-risk haplotypes had significantly increased odds of GSI, SRM and SRSR (GSI: OR 2.49 [95% CI 1.19, 4.88], p = 0.01; SRM: OR 2.97 [95% CI 1.32, 6.0], p = 0.004; SRSR: OR 5.17 [95% CI 1.65, 13.7], p = 0.001). In contrast, the association between Val174Ala alone and risk of any phenotype of intolerance among simvastatin users did not reach statistical significance. Among atorvastatin users, individuals with high-risk haplotypes had increased odds of GSI than those with low-risk haplotypes (OR 2.49 [95% CI 1.19, 4.88], p = 0.01). The association between risk haplotypes and SRM did not reach statistical significance among atorvastatin users (OR 2.71 [0.76, 7.67], p = 0.08). No association was found between Val174Ala and GSI, SRM or SRSR, nor between risk haplotypes and SRSR in atorvastatin users (see Figure 3 and Supplementary Table 5). Sample size for other statin types was insufficient for subgroup analysis.

**Fig. 3.**
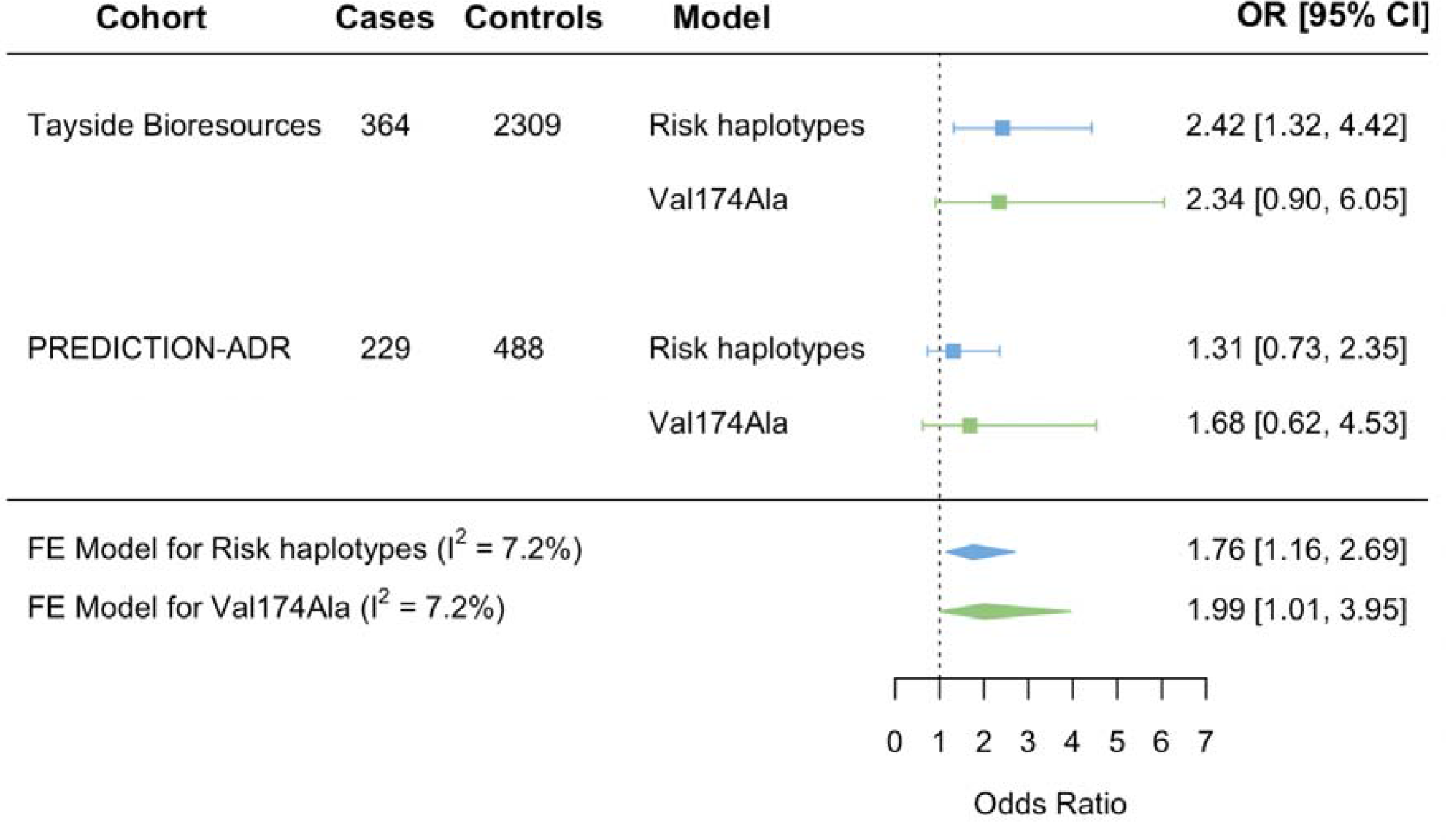
Meta-analysis of findings from Tayside Bioresources and PREDICTION-ADR. RH: Risk Haplotypes. Forest plot showing combined results from a fixed-effects meta-analysis of the association between RH and Val174Ala to statin intolerance in Tayside Bioresources and PREDICTION-ADR Consortium.

#### Meta-analysis shows risk haplotypes provide more reliable estimates of ADRs

While the association between *SLCO1B1* (either Val174Ala and risk haplotypes) and statin intolerance was not significant in the PREDICTION-ADR cohort (Fig. 3), a meta-analysis with Tayside Bioresources shows that the risk haplotypes model produces a more precise estimate of statin intolerance (OR_meta-analysis_ 1.76 [95% CI 1.16, 2.69]) than the Val174Ala genotype alone (OR_meta-analysis_ 1.99 [95% CI 1.01, 3.95]) (Fig. 3).

#### Risk haplotypes are more reliably associated with intolerance-free survival

The genetic effect of *SLCO1B1* on statin intolerance has previously been shown to be dose-dependent, consistent with the pharmacokinetic implications of the underlying causal mechanism. We therefore studied the hazards of statin intolerance by stratifying individuals by equivalent dose (<40 mg or ≥40 mg of simvastatin or equivalent dose of another statin). Val174Ala genotype and *SLCO1B1* haplotypes showed non-significant hazard ratios for all phenotypes of statin intolerance in individuals on a lower dose (see Supplementary 5: Low statin dose).

In individuals on higher statin doses (≥ 40 mg), unadjusted Kaplan-Meier curves show the cumulative risk of GSI, SRM, and SRSR according to *SLCO1B1* haplotypes (Figure 4). Adjusted hazard ratios are presented in Table 3. For all phenotypes of statin intolerance, the high-risk haplotype provided more robust estimates of time to onset of intolerance. Notably, the high-risk haplotype was able to detect an association with time to onset of SRSR 2.89 ([95% CI 1.17, 7.14], p = 0.02), whereas the Val174Ala variant alone was unable to do so.

**Figure 4.**
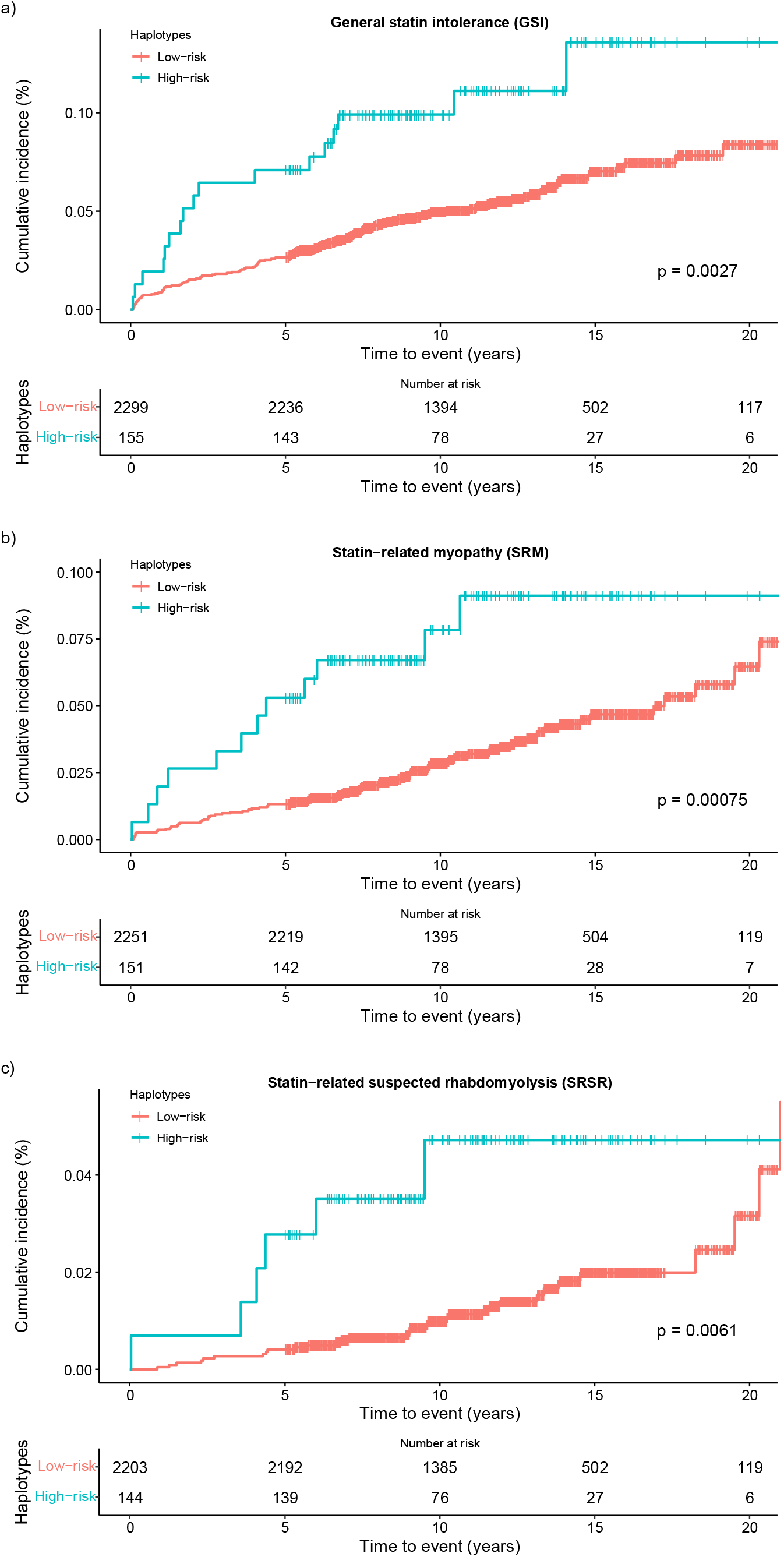
20-year intolerance-free survival by risk haplotypes in individuals on high dose. Unadjusted Kaplan-Meier curves and risk tables for cumulative incidence of a) general statin intolerance, b) statin-related myopathy, c) statin-related suspected rhabdomyolysis, in individuals on an equivalent simvastatin dose ≥40 mg, according to haplotype (blue: high-risk haplotypes; red: low-risk haplotypes).

**Table 3.** Hazards of Val174Ala and haplotypes to 20-year intolerance-free survival in individuals on higher equivalent doses (≥ 40 mg)

|  | **Ala174Ala vs.**<br>**Val174X:**<br>**HR [95% CI], p-value** | **Haplotypes:**<br>**high- vs. low-risk:**<br>**HR [95% CI], p-value** |
| --- | --- | --- |
| **GSI**<br>(n = 2,454) | 2.49 [1.09, 5.68]<br>p = 0.03 | 2.44 [1.46, 4.08]<br>p < 0.001 |
| **SRM**<br>(n = 2,402) | 3.01 [1.31, 6.94]<br>p = 0.01 | 2.51 [1.38, 4.58]<br>p = 0.003 |
| **SRSR**<br>(n = 2,347) | 2.64 [0.63, 11.1]<br>p = 0.2 | 2.89 [1.17, 7.14]<br>p = 0.02 |

CI: Confidence Interval, GSI: General statin intolerance, HR: Hazard ratios, SRS: Statin-related myopathy, SRSR: Statin-related suspected rhabdomyolysis. All models adjusted for age, gender, previous cardiovascular events, statin type and statin dose.

##### Rare variants in *SLCO1B1* are associated with statin-induced myopathy

To assess the role of rare variants, we used data from the PREDICTION-ADR consortium with 229 exome sequenced clinically adjusted cases of SRM and 488 controls. Five rare variants in *SLCO1B1* passed quality control. Their association with SRM is represented in Table 4. To analyse the burden of these variants, we performed SKAT-O analyses. The variants were found to be significantly associated with statin-induced myopathy (p = 0.02).

**Table 4.** Univariate effect of exome-sequenced rare variants on clinically adjudicated statin-induced myopathy.

| **Position (rsID)** | **Amino acid substitution** | **MAF** | **Effect** | **ORs** | **p - value** | **Directionality of effect** |
| --- | --- | --- | --- | --- | --- | --- |
| 12:21294546:C>A<br>(rs778214174) | Ala13Glu | $2.489 \times 10^{-5}$ | -1.95 | 0.14 | 0.0222 | Protective |
| 12:21325651:C>T<br>(rs769900186) | Ser51Phe | $4.956 \times 10^{-5}$ | -1.19 | 0.30 | 0.157 | Protective |
| 12:21327601:T>C<br>(rs200227560) | Ile106Thr | 0.0006932 | -0.0666 | 0.94 | 0.934 | Protective |
| 12:21331930:G>A<br>(rs147421160) | Val235Met | $3.3 \times 10^{-5}$ | -0.930 | 0.39 | 0.273 | Protective |
| 12:21358965:A>G<br>(rs74064213) | Ile499Val | 0.002482 | 0.214 | 1.24 | 0.812 | Deleterious |

These results are from the PREDICTION-ADR Consortium. Models are adjusted for age, sex, and batch effects.

## Discussion

### Summary of main findings

We provide compelling evidence from over 15,000 statin-treated individuals that assessing for the presence of GoF *SLCO1B1* variants (Leu643Phe, Asn130Asp, Pro155Thr), in addition to currently tested Val174Ala, allows for the characterization of high-risk haplotypes (RH: *SLCO1B1*\*5/*5 or *SLCO1B1*\*1a/*5). We observe that RH have a more robust association with three defined phenotypes of statin intolerance (general statin intolerance, statin-related myopathy, and suspected rhabdomyolysis) compared to the Val174Ala variant. Additionally, with risk haplotypes we observe a biological gradient of increasing effect sizes with increasing severity of statin intolerance phenotypes, especially amongst simvastatin users. This biological gradient is not observed for the Val174Ala variant. Furthermore, AIC model selection criteria show that the risk haplotypes model is a better fit for predicting statin intolerance than the Val174Ala variant alone. In a meta-analysis across our cohort and PREDICTION-ADR, we observe more robust associations of the risk haplotype with statin intolerance compared to the Val174Ala variant. Finally, for those requiring high-dose statin therapy, we observed a stronger association between high-risk haplotypes and the time to onset of general statin intolerance, statin-induced myopathy and rhabdomyolysis compared to Val174Ala. While rare variants in *SLCO1B1* have an effect on statin intolerance, this effect is modest likely due to sample size and therefore not actionable given the evidence available.

### Limitations

A limitation of this study is the lack of direct phenotype replication. Statin intolerance is a complex secondary phenotype and is challenging to define when using real-world epidemiological data. This is because it requires biochemical data alongside prescribing records and/or physicians’ notes. As such, physician-recorded adverse reactions to statins are limited and researchers rely on altered prescribing patterns and raised CK levels to identify statin intolerant individuals. These factors limit the cohorts in which the clinical phenotype of statin intolerance can be defined. Furthermore, the availability of genomic biobanks alongside such clinical data further limits study population availability. Indeed, replicating our phenotype of statin intolerance in other cohorts was a challenge. The UK Biobank is a large bioresource with genetic and clinical data; however, the only available CK testing were ordered from the primary care setting, rather than from outpatient hospital settings, such as a cardiology clinics, where cases of intolerance might be referred. Furthermore, CK testing was available for less than 50% of the UKBB cohort. This resulted in having CK measurements for only a fraction of statin users, and unusually low CK levels when compared to literature and to our discovery cohort^29^. These artefacts of the data prevented us from using the UK Biobank as a replication cohort. Further exploration of UKB laboratory data for CK is provided in Supplementary methods 4: UK Biobank cohort.

While the lack of extensive replication is a limitation, our observations are consistent with expected biological effects given the established pharmacological association between OATP1B1 and statin pharmacokinetics^15^ and are therefore less likely to be a result of type 1 error. Furthermore, they are consistent with and build upon the latest CPIC guidelines on the incorporation of gain-of-function variants to characterise OATP1B1 function and consequences for statin ADRs.

Another consideration is the exclusion of rare variants from risk haplotypes^30^. Rare variants are likely to have penetrant effects on the functional activity of the OATP1B1 transporter. However, the development of a polygenic risk score combining common and rare variants is less applicable in clinical practice since rare variants can be reliably detected only in sequenced data, which are not as readily available. Rare variants tend to be causal and act independently, however, given the frequency of statin intolerance, this is unlikely to be driven by rare variants. Furthermore, by the nature of rare variants, it is not likely that they will be in linkage with or part of haplotypes with common variants. For this reason, while we demonstrate modest effects of some rare variants, we decided against including rare variants in the risk haplotypes. Additionally, characterising common variants is more likely to result in population-level pharmacogenetic testing.

### Strengths

The main hypothesis of this study builds on the known robust role of OATP1B1 in the development of statin intolerance. Strong and consistent evidence exists on the effects of Val174Ala on statin pharmacokinetics, highlighting the importance of the functional activity of this receptor in the development of muscle symptoms. In this study, we considered all non-synonymous *SLCO1B1* variants with a MAF ≥ 0.05 to identify single- and multiple-SNPs haplotypes associated to a higher risk of statin intolerance.

By stratifying by first prescribed statin, we show that our risk haplotypes model was associated to the risk of general intolerance in both simvastatin and atorvastatin users. In comparison, no significant association could be described for the Val174Ala model for atorvastatin users. Single-dose studies suggest that *SLCO1B1* variants have a weaker, but detectable, effect on the pharmacokinetics of atorvastatin^10,16^. However, clinical trials and observational studies have failed to find an association between Val174Ala and myopathy^31,32^. These studies were likely underpowered to detect statistical significance in atorvastatin users. Better results were achieved by pooling association studies on Val174Ala and myopathy, to find an odds ratio of 2.0 per C allele in atorvastatin users^33^. That our risk haplotypes was significantly associated with intolerance also in atorvastatin users, despite a relatively small sample size (especially when compared to the statistical power of a meta-analysis), is demonstration of the specificity and biological plausibility of the effect. Even so, the odds of myopathy were considerably higher in simvastatin users than in atorvastatin users, further proof of the differential selectivity of the OATP1B1 transporter for the hepatic uptake of statins.

Another strength of this study lies in the definition of the phenotypes of intolerance. Validation against the outcome of cardiovascular events ensures robustness of the phenotypes of intolerance^20^. Particular attention was paid to the development of the control phenotype. To properly identify statin tolerant individuals, strict criteria based on laboratory and prescribing patterns were followed. Furthermore, the use of real-world longitudinal data meant that the median on-statin follow-up time was 11 (± 6) years, almost four times the average follow-up time in clinical trials.

A final strength of this study is the meta-analysis with PREDICTION-ADR which was performed in clinically adjudicated cases of statin-induced myopathy and using exome sequencing data. This confirmed our finding of a more robust association of statin intolerance with risk haplotypes compared with Val174Ala.

### Generalizability

In our study cohort, simvastatin and atorvastatin were the main drugs prescribed at the start of statin therapy (63% and 20%, respectively). While this limits the generalizability of our findings to other statins, it also reflects real-world prescription practices, particularly in the United Kingdom.

Evidence for the effect of *SLCO1B1* on rosuvastatin-associated myopathy risk is less convincing, with both pharmacokinetic and clinical studies failing to find a robust association. The small number of participants on rosuvastatin in our cohort does not allow conclusions to be drawn in this subgroup. We believe that high-quality evidence should be obtained on this specific drug, since rosuvastatin may be the optimal alternative statin in carriers of high-risk *SLCO1B1* haplotypes, especially in light of the favourable cost-effectiveness of this drug^34^.

Replicating these findings in a randomized clinical trial would allow the best quality evidence for clinical implementation; however, it would be challenging and expensive to run. Recruit-by-genotype pharmacokinetic studies would be a cost-effective alternative able to provide indirect biological evidence of these findings.

### Interpretation

Mechanistically, the association of *SLCO1B1* variants and the risk of myopathy seems straightforward. Loss of function variants disrupt the physiological function of the OATP1B1 transporter, leading to increased plasma concentration of the drug and in turn to a higher likelihood of adverse effects at the musculoskeletal level.

Our hypothesis was that the presence of protective variants might lower the risk of myopathy in statin users. By performing a comprehensive characterization of all common *SLCO1B1* exonic variants, we showed that those carrying low-risk haplotypes had significantly lower odds of myopathy than those carrying high-risk haplotypes. Interestingly, among those with low-risk haplotypes, 24% carried one copy of Val174Ala. This means that testing for Val174Ala alone would misclassify around a fourth of statin users as being at higher risk than they really are.

Of note is also the finding that Val174Ala and risk haplotypes are associated with time of statin intolerance in statin users on higher doses. This is consistent with the supposed underlying causal mechanism of the genetic effect and is in line with previous evidence on the correlation between statin dosage and risk of intolerance.

## Conclusion

In this study we show that accounting for common *SLCO1B1* gain-of-function variants provides more reliable estimates of risk and time to statin intolerance than Val174Ala testing alone. Consistent with the literature we find that impaired influx transporter function is associated with a higher risk of statin intolerance.

## Data Availability

GoDARTS and SHARE data are available subject to approval by the Tayside data access committee and can be made by contacting the corresponding authors.
PREDICTION-ADR data are available subject to approval and can be made by contacting the corresponding authors.
UK Biobank data are available via application directly to the UK Biobank, https://www.ukbiobank.ac.uk

## Acknowledgments

The authors are grateful to all study participants in the GoDARTS, SHARE, PREDICTION-ADR Consortium and UK Biobank cohort and to the respective teams.

## Ethics Statement

GoDARTS and SHARE studies were reviewed and approved by Tayside Medical Ethics Committee 053/04 and East of Scotland Ethics committee NHS REC 13/ES/0020. Multiple EU Research Ethics Committees (Tayside, Scotland; North West England, UK: Sefton, UK) approved the PREDICTION-ADR study.

UK Biobank has approval from the North West Multi-centre Research Ethics Committee (MREC) as a Research Tissue Bank (RTB) approval, REC reference: 21/NW/0157, IRAS project ID: 299116.

