## Supplementary data for "Common and rare variants in *SLCO1B1* are associated with statin intolerance"

##### Supplementary methods 1: Genetic data platforms in Tayside Bioresources

Genotype data for rs4149056, rs2306283, rs11045819, and rs34671512 were available from six platforms: Affymetrix (6.0 Affymetrix, Santa Clara), Illumina HumanOmniExpress -12VI platform, Illumina Infinium Custom GWAS chip, Illumina Infinium Global Screening Array (Illumina, San Diego), Human Exome-12 VI_A_chip, Illumina Cardio-Metabo Chip (Illumina Inc, San Diego, CA).

**Table 1 – Supplementary. Genetic data platforms, MAF and HWE.**

| SNPs | n | MAF | MAF reference population | HWE χ^2^ | HWE p value |
| --- | --- | --- | --- | --- | --- |
| rs2306283 | 15378 | 0.3840 | 0.4033 | 1.7261 | 0.18891 |
| Illumina GSA batch 1 | 4821 | 0.3884 |  | 1.8961 | 0.16852 |
| Illumina GSA batch 2 | 1824 | 0.3818 |  | 0.2421 | 0.62269 |
| Affymetrix | 3796 | 0.3818 |  | 0.9954 | 0.31843 |
| Illumina | 1813 | 0.3822 |  | 0.1498 | 0.69873 |
| Exome | 2221 | 0.3784 |  | 1.1576 | 0.28196 |
| Broad | 903 | 0.3909 |  | 0.0777 | 0.78044 |
| rs4149056 | 15378 | 0.1607 | 0.1589 | 0.0022 | 0.96259 |
| Illumina GSA batch 1 | 4821 | 0.1611 |  | 1.8999 | 0.16809 |
| Illumina GSA batch 2 | 1824 | 0.1576 |  | 0.6823 | 0.40880 |
| Affymetrix | 3837 | 0.1639 |  | 3.6017 | 0.05772 |
| Illumina | 1803 | 0.1646 |  | 0.2236 | 0.63631 |
| Exome | 2173 | 0.1569 |  | 0.0063 | 0.93674 |
| Broad | 914 | 0.1521 |  | 0.0488 | 0.82516 |
| MetaboChip | 6 |  |  |  |  |
| rs11045819 | 15378 | 0.1476 | 0.1605 | 0.0735 | 0.78631 |
| Illumina GSA batch 1 | 4821 | 0.1502 |  | 0.2846 | 0.59370 |
| Illumina GSA batch 2 | 1824 | 0.1518 |  | 0.1411 | 0.70719 |
| Affymetrix | 3845 | 0.1441 |  | 0.1727 | 0.67772 |
| Illumina | 1804 | 0.1508 |  | 0.8415 | 0.35897 |
| Exome | 2174 | 0.1389 |  | 0.0353 | 0.85097 |
| Broad | 910 | 0.1555 |  | 3.1248 | 0.07711 |
| rs34671512 | 15378 | 0.05218 | 0.05198 | 0.0205 | 0.88615 |
| Illumina GSA batch 1 | 4821 | 0.05020 |  | 0.0663 | 0.79680 |
| Illumina GSA batch 2 | 1824 | 0.05482 |  | 0.0547 | 0.81508 |
| Affymetrix | 3825 | 0.05460 |  | 0.0330 | 0.85585 |
| Illumina | 1805 | 0.05097 |  | 0.1125 | 0.73732 |
| Exome | 2191 | 0.05089 |  | 0.0208 | 0.88533 |
| Broad | 912 | 0.05263 |  | 2.8148 | 0.09340 |

### Supplementary methods 2: Exome sequencing for rare variants

**Library preparation and sequencing**

Sequencing of samples was performed using exome-enriched sequence data (SureSelect QXT, XT and XT2 reagents (Agilent Technologies, Wokingham, UK)). Hybridization capture enrichment of whole genome libraries was performed using the SureSelect v5 all-exon probe set, following manufacturer’s recommendations throughout (<http://www.agilent.com/cs/library/usermanuals/Public/G9681-90000.pdf>). Equimolar aliquots of 12 or 10 post-enrichment libraries (6 cases and their 6 matching controls) were pooled before sequencing using version 2 TruSeq chemistry on a Nextseq500 or Hiseq2500 (Illumina Inc., San Diego, CA, USA).

**Read mapping and variant calling**

Paired-end 150-bp sequence reads were then analysed using an in-house pipeline. The pipeline was made of BWA-MEM, Genome Analysis Toolkit (GATK) v.1.6 UnifiedGenotyper, Cutadapt, Picard v1.119 and Annovar.

**Data analysis and quality control**

Processed sequence data passed quality control if they achieved at least 20x for more than 70% on targeted bases, Ts/Tv ratio, Het/Hom ratio, and deletion/insertion ratio. Sample metrics were compared between the 3 different centres and no significant deviation of calls or samples were noted with 99.8% genotype concordance.

### Supplementary methods 3: Validation of phenotypes of intolerance

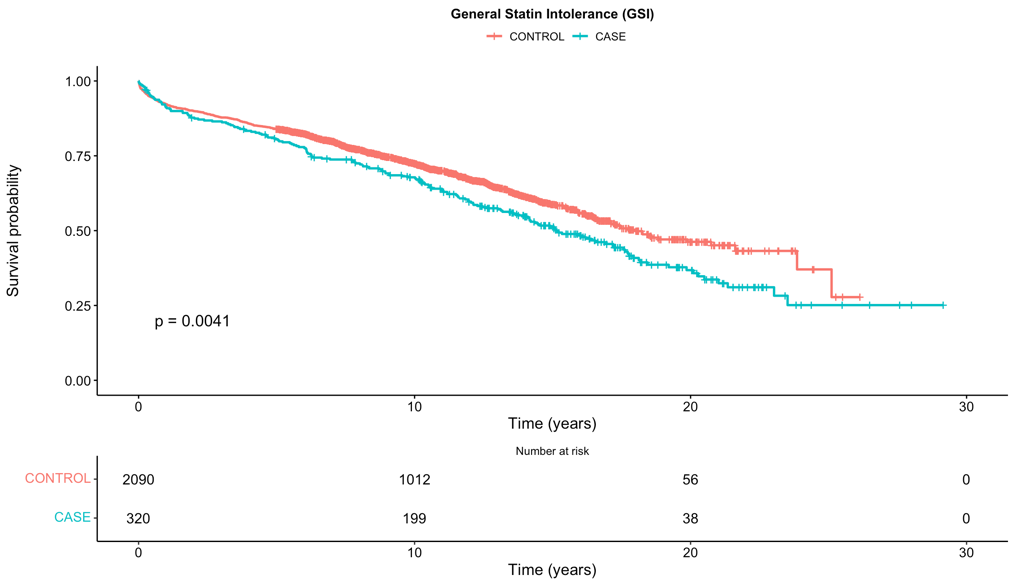
Figure 1 – Supplementary. Time to statin failure among the three phenotypes.

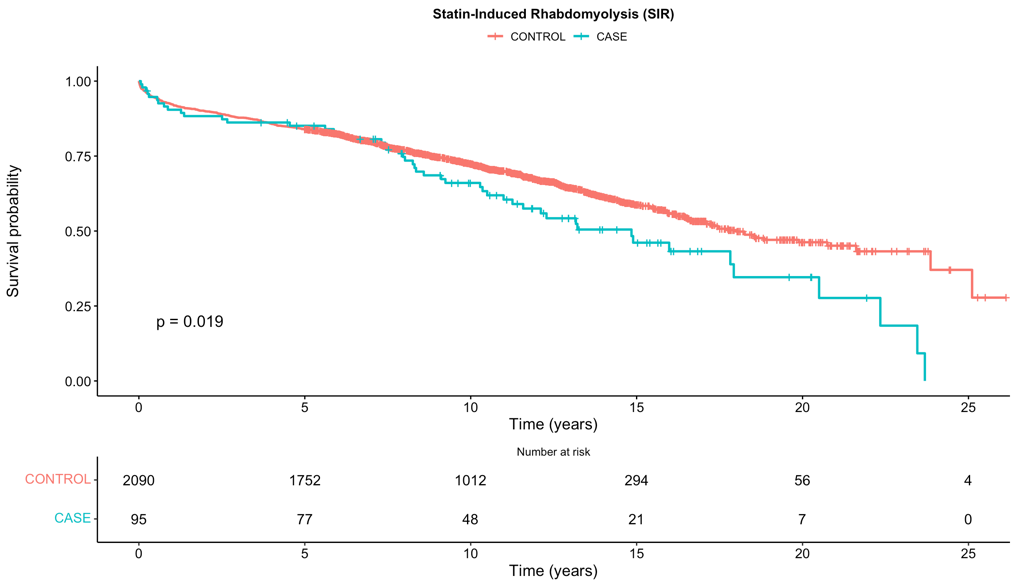

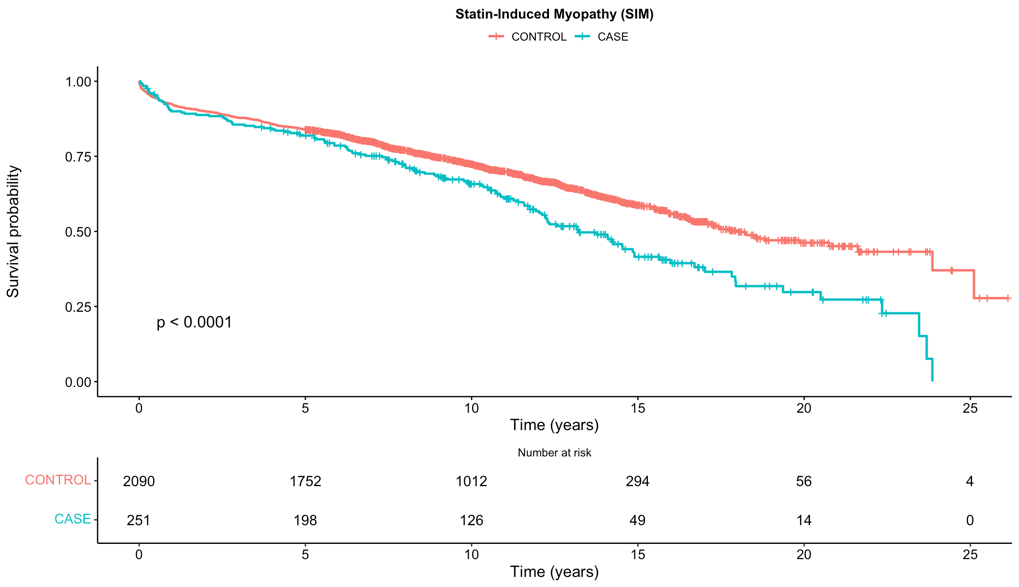

**Table 2 – Supplementary. Hazards of statin failure among the three phenotypes of intolerance.**

| **Phenotypes of intolerance** | **HR** | **95% CI** | **p value** |
| --- | --- | --- | --- |
| **GSI** | 1.39 | 1.16 – 1.68 | <0.001 |
| **SRM** | 2.18 | 1.73 – 2.74 | <0.001 |
| **SRSR** | 3.06 | 2.24 – 4.18 | <0.001 |

CI: Confidence Interval, GSI: General Statin Intolerance, HR: Hazard Ratio, SRM: Statin-related Myopathy, SRSR: Statin-related suspected rhabdomyolysis. All models adjusted for age, sex, previous cardiovascular events, statin dosage.

### Supplementary methods 4: UK Biobank cohort

From 230,000 participants for which primary care data is available, 63,433 were identified as statin users on the bases of prescription records. Of these, 61,513 have complete and plausible prescribing information recorded. One individual was excluded due to a lack of genotype information.

Creatinine kinase (CK) levels measured while on statin therapy were available for 10,847 statin users. Both average all-time CK levels and average highest CK levels were lower than expected in statin users [PMID: 23958263] and lower than the recorded values in Tayside Bioresources (median highest CK levels in UKBB 89.0 UI/L [IQR 58-129] vs median highest CK levels in discovery cohort 145 UI/L [IQR 92-234]) (Fig. 2). This is likely because laboratory data from patients referred to specialist clinics are not available in the UKBB as of the date of writing.

This resulted in two-fold higher CK levels in cases when compared to controls in UKBB, versus a three-fold increase in the Tayside Bioresources cohort. More importantly, the lack of CK measurements renders unreliable the definition of statin tolerant controls, which is based on the lack of CK testing for the required 5 years (minimum) of statin therapy. For these reasons, we decided against including findings from the UKBB in our meta-analysis results.

**Fig. 2. Comparison of CK levels between discovery cohort (Tayside Bioresources) and UK Biobank**

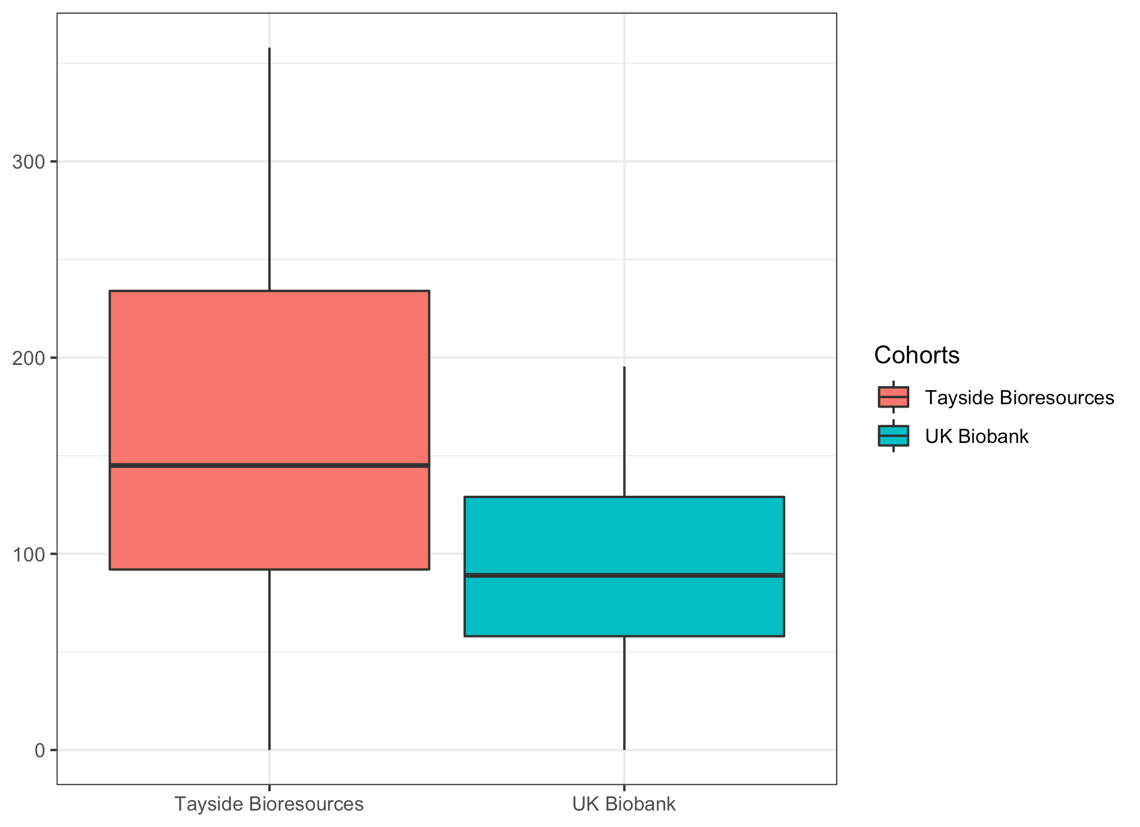

### Supplementary methods: Statin type

**Table 3 – Supplementary. Association of Val174Ala and risk haplotypes with statin intolerance by first statin type.**

| Atorvastatin |  | GSI | SIM | RDM |
| --- | --- | --- | --- | --- |
|  |  | n = 614 | n = 611 | n = 577 |
|  | Ala174Ala | OR: 2.85 [0.13, 22.1], p = 0.38 | OR: 2.81 [0.42, 11.0], p = 0.19 | 2.17e-7 [NA, 2.19e66], p = 0.994 |
|  | High-risk haplotypes | OR: 5.21 [1.26, 18.6], p = 0.01 | OR: 2.71 [0.76, 7.67], p = 0.08 | 1.44 [0.007, 7.93], p = 0.73 |
| Simvastatin |  | n = 1,867 | n = 1,828 | n = 1,708 |
|  | Ala174Ala | OR: 2.62 [0.82, 7.04], p = 0.07 | OR: 2.96 [0.86, 7.78], p = 0.051 | OR: 4.36 [0.67, 16.1], p = 0.055 |
|  | High-risk haplotypes | OR: 2.49 [1.19, 4.88], p = 0.01 | OR: 2.97 [1.32, 6.0], p = 0.004 | OR: 5.17 [1.65, 13.7], p = 0.001 |

### Supplementary methods 5: Low statin dose

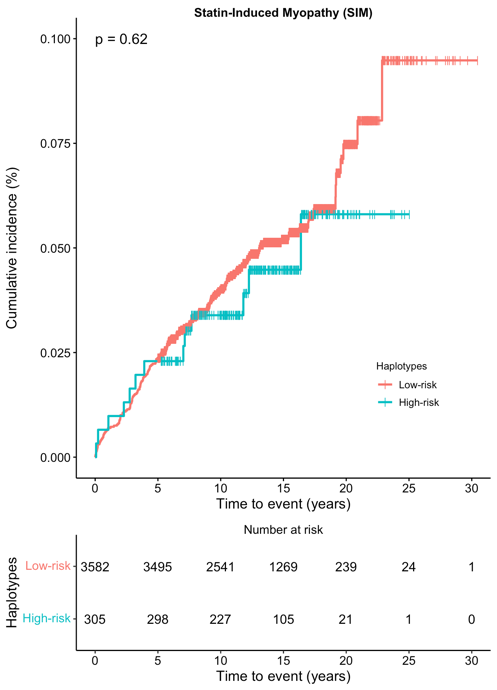

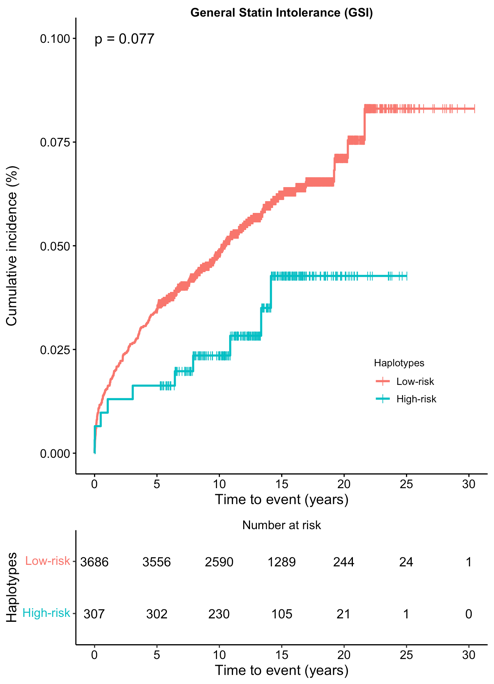

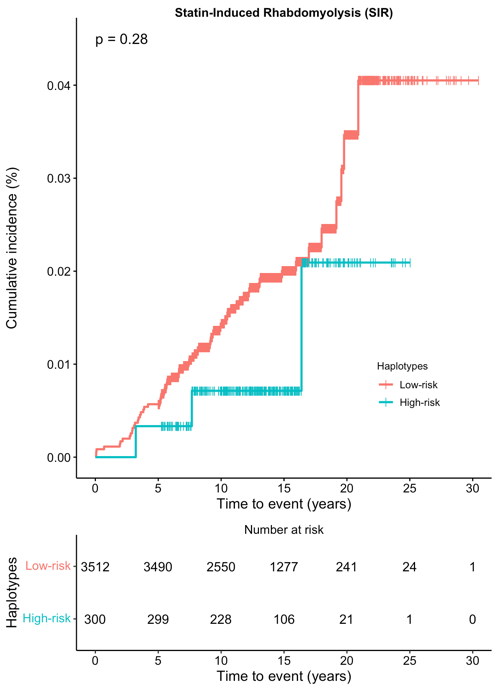
**Figure 3. Time to onset of statin intolerance among the three phenotypes of intolerance, in individuals on lower equivalent doses (<40mg).**

**Table 4 – Supplementary. Hazards of Val174Ala and haplotypes to statin intolerance in individuals on lower equivalent doses (< 40 mg).**

|  | **Ala174Ala vs.**  **Val174X:**  ***HR [95% CI], p-value*** | **Haplotypes:**  **high- vs. low-risk:**  ***HR [95% CI], p-value*** |
| --- | --- | --- |
| **GSI**  (n = 3,993) | 0.63 [0.23, 1.69]  p = 0.4 | 0.60 [0.31, 1.11]  p = 0.10 |
| **SIM**  (n = 3,887) | 0.68 [0.25, 1.83]  p = 0.7 | 0.88 [0.50, 1.55]  p = 0.7 |
| **RDM**  (n = 3,812) | 1.40 [0.44, 4.46]  p = 0.3 | 1.40 [0.44, 4.46]  p = 0.3 |
